# Preclinical mobility limitation outcomes in rehabilitation interventions for middle-aged and older adults population: a scoping review protocol

**DOI:** 10.1101/2022.10.22.22280644

**Authors:** Aiping Lai, Ashley Morgan, Julie Richardson, Lauren Griffith, Ayse Kuspinar, Jenna Smith-Turchyn

## Abstract

**Objectives:** This scoping review aims to understand the extent of evidence regarding preclinical mobility limitation (PCML) intervention studies that have been implemented or planned in middle-aged and older adult populations.

**Introduction:** Individuals with PCML are at a high risk of future functional loss and progression to disability. An overview of studies undertaken on this emerging topic is now due.

**Inclusion criteria:** Rehabilitation intervention studies that measured PCML outcomes or assessed individuals at the PCML stage will be included. Studies will be considered if the participants are middle-aged (45-64yrs) or older adults (≥ 65yrs) in any setting, including community-dwelling, hospital discharges, or institutional settings.

**Methods:** Seven databases (MEDLINE, EMBASE, AMED, PsycINFO, CINAHL, Web of Science and Cochrane CENTRAL) were searched to locate relevant published and unpublished intervention studies (English evidence from inception onwards). The search strategy will be generated using the PCC framework (population, concept, and context) and refined after consulting with a McMaster research librarian. In addition, a manual search from the reference list of retrieved papers and review articles will also be performed. Two reviewers will use predefined inclusion/exclusion criteria to independently review titles, abstracts, and full texts of potential articles. Any disagreements on study selection will be resolved by discussion or consensus involving a third reviewer. Data will be collected and reported using a predefined data extraction chart and described using qualitative content analysis.

## Introduction

Mobility is defined as the ability to move oneself (independently or by using assistive devices or transportation) around their environment^1–2^. Mobility is a key physical ability that influences the quality of life as a strong prognostic indicator of disability^3^. A mobility limitation is usually the first area in which disability occurs. Twenty-four percent of older Canadians aged 65 to 74 experienced mobility disability, and this percentage increases as they grow older. Forty-one percent of older adults aged 75 to 84 years and 61 percent of those aged 85 years and over experience mobility disability^4^. The trajectory of mobility loss with aging is commonly described as a descending curve with a steeper decline shown late in life^3^. However, age-related impairments in mobility-associated physiological systems can be compensated by changing the manner or the frequency of doing a mobility task^5^. An overt mobility limitation only manifests when the severity of mobility loss becomes too high to be compensated^3^. The intermediate stage, which occurs with progressive loss of function and precedes the onset of disability, was referred to as “preclinical disability”^6^, “preclinical mobility disability”^7^, or “preclinical mobility limitation” (PCML)^8^. The concept of preclinical disability was first introduced by Fried and colleagues^6^. They defined it as a functional stage where people can still complete certain tasks but change the frequency or modify the way of doing the tasks. They linked the progressive disability to an iceberg and described the stage of PCML as a submerged portion of the iceberg. Only the tip of the iceberg is visible, representing individuals can no longer compensate successfully for their loss of physical function.

Before significant mobility and functional independence losses, there may be interventions that can improve recovery likelihood and functional levels^9^. Individuals in the PCML stage may represent an optimal group to receive disability prevention interventions, as they are likely to be responsive to altered risk factors^10^. In the primary prevention of disability among older adults, intervening at the PCML stage might be the most effective strategy to reduce the burden of disability in this population. However, studies have provided inconclusive results. For example, an RCT study compared older adults with preclinical gait dysfunction in a motor learning program incorporating goal-oriented stepping and walking with those in a standard program employing endurance training by treadmill walking^11^. Both groups improved walking endurance, but older adults in the motor learning group demonstrated more improvements in mobility performance parameters (such as gait speed and motor skill)^11^. Yet, Day and colleagues investigated a group of community-dwelling older adults with PCML in a Tai Chi group and seated flexibility exercise group for 24 weeks and found little change in impairments or functional limitations ^12^. Many factors may contribute to these conflicting findings, such as tools used to identify the stage of PCML, eligibility criteria for participants, and interventions selected. A preliminary search also showed that the outcomes measuring changes of PCML vary among studies^12–14^. Some used self-reported measures, such as the preclinical disability screening tool by Fried et al.^15^ or the Preclinical mobility disability scale by Manty et al.^8^, while others debate whether performance-based measures can identify more limitations in physical function than self-report measures^11^. In addition, the characteristics of the sample recruited (e.g. age, gender, cognitive level, etc.) and the variation in intervention used (e.g. type, frequency, length, intensity and delivery professions, etc.) may also contribute to the variation in results.

However, there was a lack of resource mapping evidence in relation to these concepts, which leads to the uncertainty of whether or not it would be helpful to conduct a systematic review. In addition, determination of the key items required when undertaking a systematic review, such as identifying precise research questions and defining suitable inclusion criteria.

Therefore, synthesizing the evidence currently available in PCML interventions will help determine what these interventions are, what type of interventions are associated with various outcomes and future directions for interventional research. A preliminary search in MEDLINE, the Cochrane Database of Systematic Reviews, Joanna Briggs Institute (JBI) Evidence Synthesis, and PROSPERO was conducted. No current or ongoing systematic or scoping reviews on this topic were identified. The purpose of this scoping review is to provide a comprehensive understanding of PCML intervention studies in middle-aged and older adults that have been tested or planned, map how they have been conducted and reported, identify the knowledge gaps in current literature and make recommendations about future research direction in the interventional study in PCML.

### Review questions

- What outcomes are used to measure PCML changes in the intervention studies? What other outcomes are reported in association with PCML outcomes?
- What types of rehabilitation interventions are used to change PCML outcomes in middle-aged and older adults?
- What are the characteristics of the baseline samples included in the PCML intervention studies (e.g., participants’ baseline characteristics, eligibility criteria, PCML stage assessment)?

### Eligibility criteria

#### Participants

Studies will be considered for inclusion in this review if there are PCML outcomes reported or if participants are identified as at the PCML stage. All the participants in the included studies will be middle-aged (45-64yrs) or older adults (65yrs or over).

#### Concept

The concepts of the current review are rehabilitation intervention studies where PCML outcomes were reported or people who reported functional changes consistent with the PCML stage received a rehabilitation intervention. A rehabilitation intervention was defined as any non-surgical or non-pharmacological intervention^16^. Studies using a rehabilitation intervention as a single-component intervention or as a part of a multifaceted intervention regardless of frequency, intensity, length, and who delivered them will be included. Measurements of PCML stage and outcomes could be either self-reported (e.g. Fried task modification and disability scale, Manty scale, etc.) or physical performance measures (e.g. CS-PFP10, gait speed, etc.) as long as the term of PCML or synonym (e.g. preclinical disability, subclinical disability, etc.) as indicated explicitly in the studies.

#### Context

The present review will consider the studies conducted in any setting, including community-dwelling, hospital settings (e.g. emergency room, hospital admission, rehab center, etc.), or institutional settings (e.g. long-term care, supported housing, etc.).

#### Types of study to be included

This scoping review will consider experimental and quasi-experimental study designs, including randomized controlled trials (RCTs), non-randomized controlled trials (NRS), controlled clinical trials, pre-post studies, and interrupted time series studies. In addition, uncontrolled longitudinal studies, case studies, registered trials, and protocols will also be considered for inclusion. Review papers will also be used to identify original studies. Two reviewers will scrutinize the reference lists of eligible review papers to determine additional studies for review. Abstract-only publications will not be considered for inclusion.

## Method

The first methodological framework for scoping reviews was developed by Arksey and O’Malley^17^. It then further developed over the years and was extended by Levac et al.^18^. The proposed scoping review will be conducted in accordance with the JBI methodology for scoping reviews which a working group developed from JBI and the JBI Collaboration (JBIC)^19–20^.

### Search strategy

A search will be conducted in seven databases (MEDLINE, EMBASE, AMED, PsycINFO, CINAHL, Web of Science and Cochrane CENTRAL) to locate relevant intervention studies (English evidence from inception to March 19, 2022). The search strategy will be generated using the PCC framework (Population, Concept, Context) and refined after consulting with a McMaster research librarian. The text words included in the titles and abstracts of relevant papers and the index terms used to describe the papers were used to develop a complete search strategy for Ovid MEDLINE (see Appendix I). The search strategy will be adapted for each database, including all identified keywords and index terms. In addition, a manual search from the reference list of retrieved papers and review articles will also be performed for additional studies.

### Study selection

After the search is completed, all identified citations will be collated and uploaded into Covidence (Veritas Health Innovation, Melbourne, Australia). Duplicate records will be removed in Covidence. Before screening all retrieved articles, two reviewers will conduct a pilot test of the eligibility criteria on the first 20 articles in alphabetical order of the author’s last name. The eligibility criteria will be modified as necessary for clarity based on the experience with the pilot test. After the pilot test, two reviewers will independently review titles, abstracts, and full texts of potential articles. Reasons for excluding sources of evidence in full text that do not meet the inclusion criteria will be recorded and reported in the scoping review. Any disagreements about study selection will be resolved by discussion or consensus involving a third reviewer. The research results will be reported in full in the final scoping review in JBI Preferred Reporting Items for Systematic Reviews and Meta-Analyses for scoping reviews (PRISMA-ScR) flow diagram^21^.

### Data extraction

Two reviewers will conduct the data extraction to reduce errors and bias. A data extraction table will be developed, pilot-tested, revised and finalized to ensure all records are extracted with the same criteria. The data extracted will include the author’s last name, publication year, types of study, sample size, participants’ characteristics (age, gender, setting) by groups (intervention vs. comparator), and specific details about the interventions (e.g. content, length and frequency, single or multifaceted, outcomes reported). A draft extraction form is provided (see Appendix II). Any modifications to the data extraction form will be detailed in the review. If appropriate, authors of articles will be contacted to request missing or additional data.

### Data analysis and presentation

The extracted data will be presented in tabular form that aligns with the research questions of this scoping review. Interventions will be presented by listing type, content, length, intensity, frequency, and whether it is delivered as a single method or part of a multifaceted approach. Participants’ characteristics will be presented by listing age, gender (% of female and % of male), % of participants identified as at the stage of PCML and the measurements used. Self-reported or physical performance measured PCML outcomes and other outcomes reported in the studies will also be presented. A narrative summary will accompany the tabulated results and describe how the results relate to the review questions.

## Data Availability

All data produced in the present study are available upon reasonable request to the authors

### Appendices

#### Appendix I: Search strategy

OVID MEDLINE (Search conducted on March 19, 2022)

|  |  |  |
| --- | --- | --- |
| 1 | exp Middle Aged/ | 4669400 |
| 2 | exp Aged/ or exp "Aged, 80 and over"/ | 1532743 |
| 3 | ((old* or elder* or geriatric*) adj5 (adult* or patient* or person* or people* or men or man or woman or women)).mp. [mp=title, abstract, original title, name of substance word, subject heading word, floating sub-heading word, keyword heading word, organism supplementary concept word, protocol supplementary concept word, rare disease supplementary concept word, unique identifier, synonyms] | 888364 |
| 4 | middle aged.mp. | 4684594 |
| 5 | 1 or 2 or 3 or 4 | 5491201 |
| 6 | ((preclinical* or pre-clinical or subclinical* or sub-clinical or incipient* or perceived or early) adj3 (mobility or function* or physical) adj3 (disab* or declin* or limit* or deficit* or dysfunction* or impair* or difficult* or deteriorat* or modif*)).mp. [mp=title, abstract, original title, name of substance word, subject heading word, floating sub-heading word, keyword heading word, organism supplementary concept word, protocol supplementary concept word, rare disease supplementary concept word, unique identifier, synonyms] | 2008 |
| 7 | ((preclinical* or pre-clinical or subclinical* or sub-clinical) adj (disab* or declin* or limit* or deficit* or dysfunction* or impair* or difficult* or deteriorat* or modif*)).mp. [mp=title, abstract, original title, name of substance word, subject heading word, floating sub-heading word, keyword heading word, organism supplementary concept word, protocol supplementary concept word, rare disease supplementary concept word, unique identifier, synonyms] | 622 |
| 8 | 6 or 10 | 2592 |
| 9 | 5 and 11 | 1247 |
| 10 | limit 12 to the English language | 1191 |

#### Appendix II: Data extraction instrument

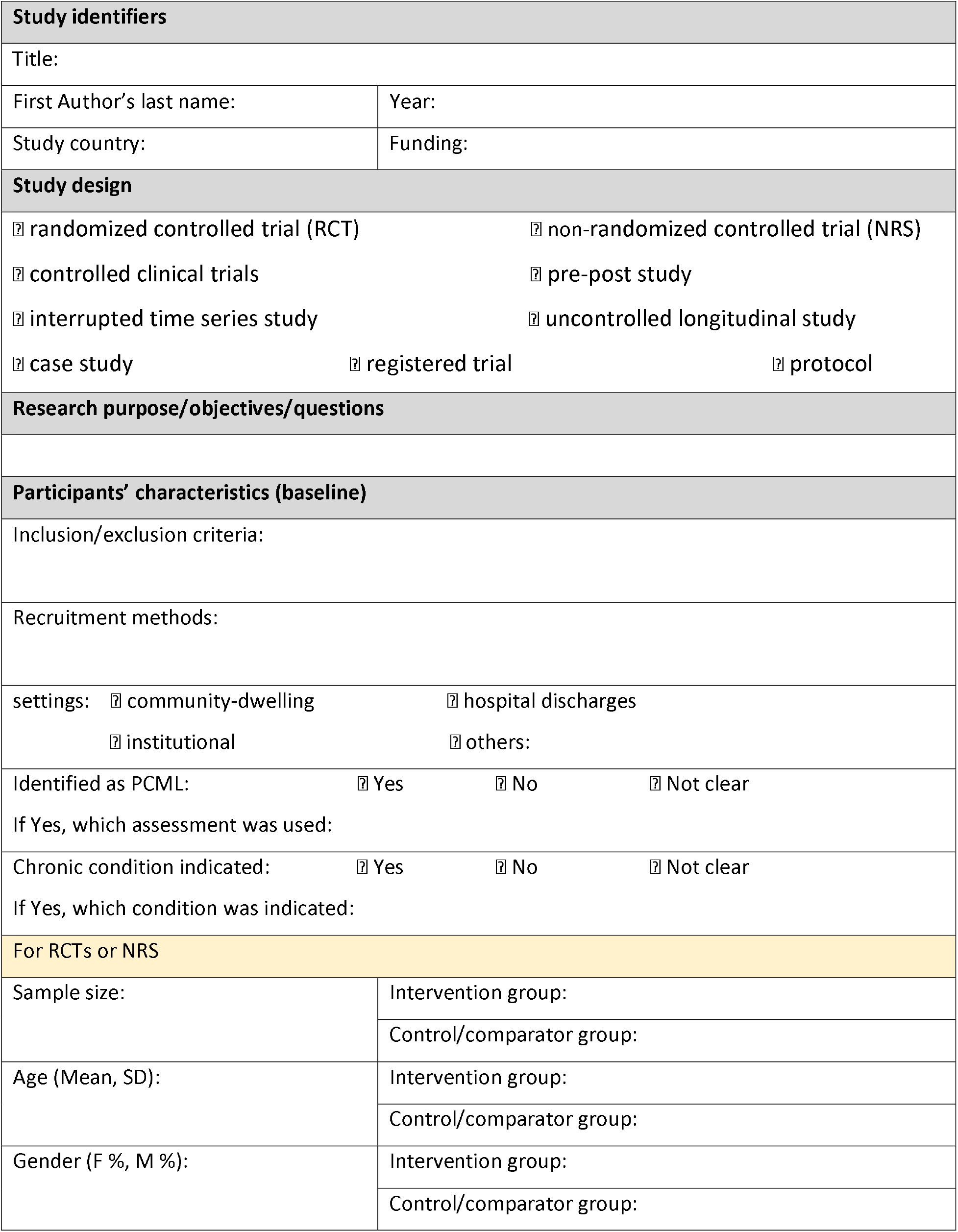
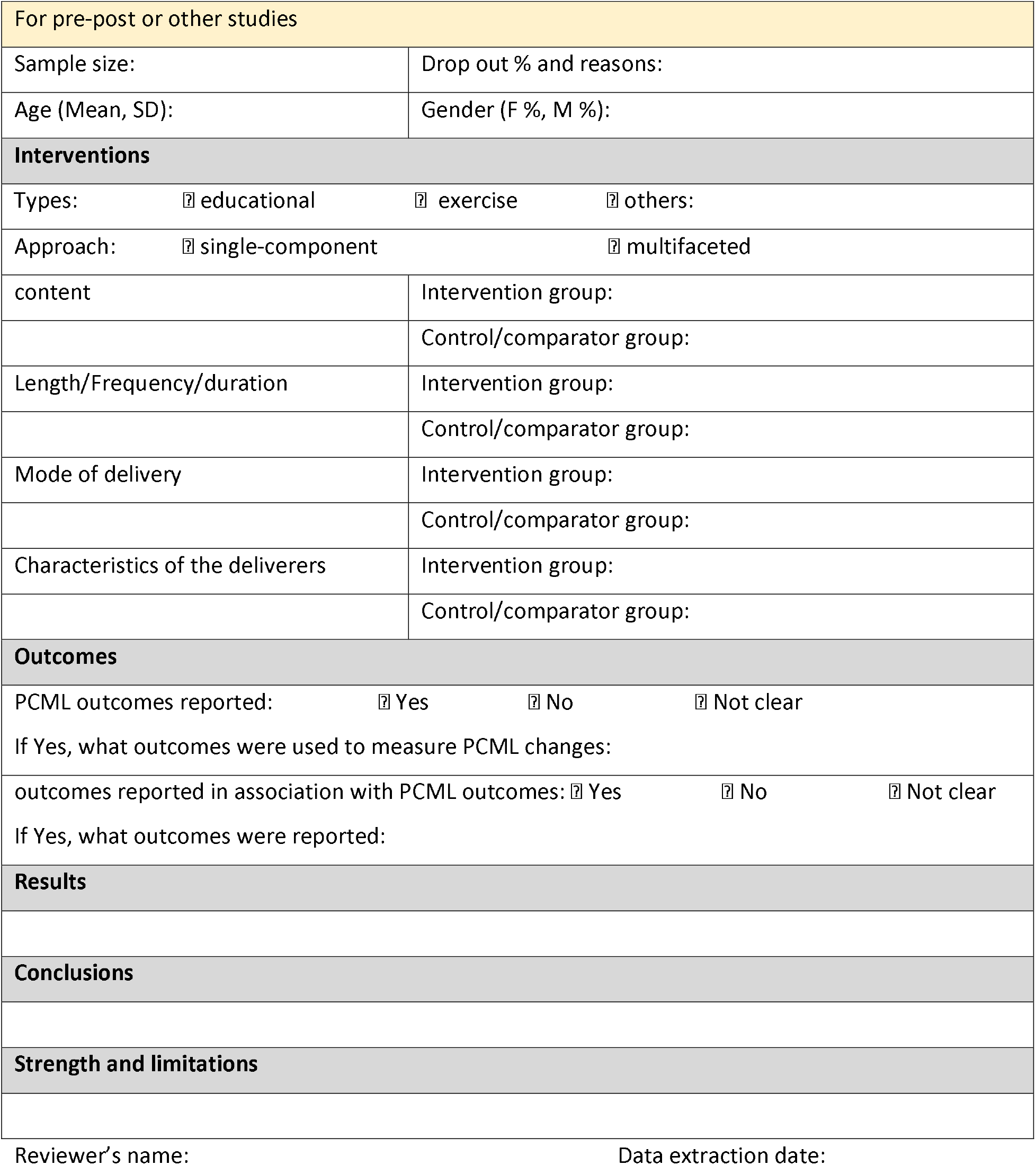

| Study identifiers |  |  |  |
| --- | --- | --- | --- |
| Title: |  |  |  |
| First Author's last name: |  | Year: |  |
| Study country: |  | Funding: |  |
| Study design |  |  |  |
| ☐ randomized controlled trial (RCT) |  | ☐ non-randomized controlled trial (NRS) |  |
| ☐ controlled clinical trials |  | ☐ pre-post study |  |
| ☐ interrupted time series study |  | ☐ uncontrolled longitudinal study |  |
| ☐ case study | ☐ registered trial | ☐ protocol |  |
| Research purpose/objectives/questions |  |  |  |
| Participants' characteristics (baseline) |  |  |  |
| Inclusion/exclusion criteria: |  |  |  |
| Recruitment methods: |  |  |  |
| settings: ☐ community-dwelling |  | ☐ hospital discharges |  |
| ☐ institutional |  | ☐ others: |  |
| Identified as PCML: | ☐ Yes | ☐ No | ☐ Not clear |
| If Yes, which assessment was used: |  |  |  |
| Chronic condition indicated: | ☐ Yes | ☐ No | ☐ Not clear |
| If Yes, which condition was indicated: |  |  |  |
| For RCTs or NRS |  |  |  |
| Sample size: | Intervention group: |  |  |
|  | Control/comparator group: |  |  |
| Age (Mean, SD): | Intervention group: |  |  |
|  | Control/comparator group: |  |  |
| Gender (F %, M %): | Intervention group: |  |  |
|  | Control/comparator group: |  |  |

|  |  |
| --- | --- |
| For pre-post or other studies |  |
| Sample size: | Drop out % and reasons: |
| Age (Mean, SD): | Gender (F %, M %): |
| **Interventions** |  |
| Types: ☐ educational ☐ exercise ☐ others: |  |
| Approach: ☐ single-component ☐ multifaceted |  |
| content | Intervention group: |
|  | Control/comparator group: |
| Length/Frequency/duration | Intervention group: |
|  | Control/comparator group: |
| Mode of delivery | Intervention group: |
|  | Control/comparator group: |
| Characteristics of the deliverers | Intervention group: |
|  | Control/comparator group: |
| **Outcomes** |  |
| PCML outcomes reported: ☐ Yes ☐ No ☐ Not clear |  |
| If Yes, what outcomes were used to measure PCML changes: |  |
| outcomes reported in association with PCML outcomes: ☐ Yes ☐ No ☐ Not clear |  |
| If Yes, what outcomes were reported: |  |
| **Results** |  |
| **Conclusions** |  |
| **Strength and limitations** |  |

Reviewer's name:
Data extraction date:

